# DIETARY PRACTICES AND NUTRITION STATUS OF PREGNANT ADOLESCENTS’ IN KAJIADO COUNTY, KENYA

**DOI:** 10.64898/2026.07.28.26359156

**Authors:** Caroline Wambui Wandu, Joseph Kobia, Dorcus Mbithe David-Kigaru

## Abstract

Kajiado County ranks eighth in the country for adolescent pregnancy, with one in five teenagers between the ages of 15 and 19 (21.8%) likely to be pregnant or have begun having children. The objectives of the study were to establish the dietary practices and nutrition status of pregnant adolescent’s in Kajiado County. The study employed a cross-sectional analytical design in a target population of 196 pregnant adolescents living in Kajiado West, Ewauso and Keekonyoike wards which were randomly selected. Qualitative data was gathered through key informant interview guides with facility nurse in charge and head teacher/principal. Dietary data was collected using a 24-hour dietary recall questionnaire.Dietary diversity was established using the minimal dietary diversity questionnaire for women of reproductive age (MDD-W). Nutrisurvey software was to analyze dietary data. Married respondent was at 41.8% (40.8 users and 44.1% non-users) of ANC with a p-value of 0.212. Dietary energy intake was 1086.82 ± 90.57kcals having RDA of 29.0% (users at 1105.72 ±95.43 and RDA 29.6%, non-users 1070.12±8.36 with RDA of 28.4%). The Minimum Dietary Diversity (MDD-W) for Women of Reproductive Age 98.4% users was 68.3% of women who utilized antenatal care (ANC) services consumed foods and beverages from 5 or more food groups on the previous day, compared to only 31.7% non-users having a p-value 0.24. Respondent having (MUAC < 22 cm) indicates potential malnutrition or nutritional risk at 31.2% (users at 30.8% and non-users 32.2%) with a p-value 0.534.Overall mean hemoglobin level is 12.35g/dl with a standard deviation of 0.93g/dl having low ranges of < 11.5g/dl (users at 30 and non-users at 12) total of 42 adolescent. These findings underscore the urgent need for targeted interventions at increasing nutrient intake and dietary diversity among pregnant adolescents towards improved nutrition status.

## Introduction

Adolescent pregnancies are believed to cause serious health, malnutrition, psychosocial and economic risks to the girls which impedes their ability to have children advance their careers. These thus does trap them in poverty cycles mostly being from low-income families and limiting their potentials and choices in life [1]. Pregnant adolescent girls can serve as a key entry point for interventions aimed at establishing and maintaining healthy eating habits throughout adulthood. Pregnancy outcomes in women of reproductive age are based on the nutrition status thus leading to the birth weight of the newborn.

In developing regions, approximately 21 million girls in this age group become mothers annually. In Kenya, adolescents significant proportion of girls aged 15-19 are pregnant with their first child, with estimates indicating about 21.8% [2].

The utilization of antenatal clinic (ANC) services is crucial for ensuring positive pregnancy outcomes and reducing maternal and neonatal mortality. Regular ANC visits provide essential health education, nutritional support, immunizations, and early detection of pregnancy-related complications. Studies have demonstrated that women who attend ANC services are more likely to receive vital interventions such as tetanus toxoid vaccinations and iron supplements, which significantly contribute to improved maternal and neonatal health [3]. Conversely, the non-use of ANC services is associated with higher risks of adverse outcomes, including preterm labor, low birth weight, and perinatal mortality. Barriers such as socio-economic challenges, cultural beliefs, and limited access to healthcare facilities contribute to the underutilization of ANC services [4]. Addressing these barriers through community education, improved healthcare infrastructure, and culturally sensitive interventions is essential for increasing ANC attendance and enhancing maternal health outcomes.

## Materials and methods

### Ethics statement

The Kenyatta University Graduate School provided research approval. Approval from Kenyatta University’s Ethical Review Committee KU/ERC/APPROVAL/VOL.1. PKU/2863/11986 and permission was issued by the National Council for Science, Technology, and Innovation (NACOSTI) NACOSTI/P/24/32720. Kajiado County; Director of Medical Services CGK/MEDICALSERVICES/01/3)VOL.11/303. Ministry of Interior and National Administration County Commissioner Kajiado (KJD/CC/ADM/45VOL.IV (87) and Director of Education KJD/C/R.3/111/85. The respondent’s parents/guardians or husbands did grant their consent, while pregnant adolescent gave ascent. There were no risk associated with this research as a medical lab technician was used in the collection of the sample at homestead. Standard operating procedures (SOPs) for collecting blood samples were used [5]. A guarantee of confidentiality was made and maintained after the study.

### Setting and research design

The study was carried out in Kajiado West, a Sub County within Kajiado County. It has a total population of 1,117,840 people. In 2016, the county’s poverty rate was 36.9%. Pastoralism, tourism, and agriculture are the main economic pursuits [9]. The resident ethical community is the Maasai. Most of the time, the sub-food county’s security is precarious, which has an impact on local households’ dietary intake and nutrition [9]. The residence ethical community is the Maasai. Kajiado West sub-county was chosen as one of the regions frequently affected by the droughts as it is arid region. In Kenya 15% of adolescents aged 15 to 19 are already pregnant while Kajiado County being 8^th^ nationally at 21.8% hence a justification of this study location [10]. The research used a cross-sectional analytical study design, which allowed for collection of both qualitative and quantitative data and testing of numerous hypotheses. It had a comparative component.

### 3.4 Study Population

The study aimed at all pregnant adolescents aged 10 to 19 years in Kajiado West Sub-County.

### Determination of Sample Size

Fisher formula [11] of cross sectional studies was used to determine the sample size as follows:

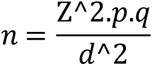

requested sample size (if the target population is greater than 10,000) p = Proportion of population (21.8% percentage of women age 15-19-year-old who have ever been pregnant in Kajiado County [10]. This prevalence would provide the greatest number of respondents for the necessary minimum sample size).

Z = Standard normal deviate (1.96) at the necessary level of confidence.

q is equal to the population minus the traits being measured (1-p)

d = Level of accuracy (0.05)

n = 1.96

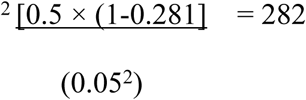

To ensure a sample size is comparable to the population, Fisher’s finite adjustment for proportionality will be used using the formula:

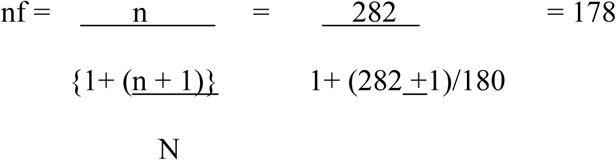

The above number had an additional 10% (18) added to account for non-response, resulting in a final sample size of 196 adolescents. This included 98 users and 98 non-user of ANC.

### Sampling procedure

The sampling was conducted using multi-stage random sampling. Kajiado West sub-county region was purposively selected due to high levels of poverty and frequent food supply shocks, ensuring the study focuses on areas significantly impacted by these issues. Random sampling was done in the 5 wards namely Ewauso, Iloodokilanik, Keekonyoike, Magadi and Mosiro where Ewauso and Keekonyoike were chosen.

Data was collected from 19 health institutions of the randomized sample of 38 health facilities from Ewauso (24) and Keekonyoike (14) wards which aided in getting records of 98 ANC pregnant adolescent users in both wards. Records of ANC pregnant adolescent users were used. A total of 210 pregnant adolescent ANC users were recovered from health facility records of which they were randomly sampled on every 2^nd^ household. The 38 health institutions linked respective schools where ANC non-users were identified. Ewauso having 22 schools and keekonyoike 21 schools.

Community Health Promotors (CHP) did also aided in identifying the ANC non-users in the wards. CHPs and records from schools identified 150 households with ANC non-users. In Keekonyoike ward, a total of 60 households were identified and 90 households in Ewauso ward. Simple random sampling was used to choose from 150 household with pregnant teens in the two wards. The first household was chosen using a table of random numbers and each n^th^ number (every 2^nd^) household was drawn to achieve 98 ANC non-users. Hemoglobin (Hb) levels were determined by analysis of blood samples taken from all participants by a competent medical lab technician.

### Methods of data collection

#### Recruitment period for this study

Data was collected at households between the 2^nd^ February 2024 and the 30^th^ May 2024.

#### Structured questionnaire

A structural administered questionnaire was at the household level and similar for both users and non-users of antenatal clinic which entailed demographic, socio-economic and socio-cultural characteristics. The pregnant adolescent respondent level and sources of nutrition knowledge was assessed in the household a set of 11 questions were asked each having 9.09 points to add to 100%. Then gave each respondent a % overall score, using the classification of <40% as low, 41-69 as moderate and >70% as high. Dietary practices and dietary intake which entailed a 24-hour dietary recall questionnaire where the respondent did recall food and drinks consumed and their quantities and preparation methods in the past 24 hours. Food plate demos and serving spoons were used to aid in amount recall, quantitative data on nutrient consumption and meal frequency was gathered. Data on dietary variety was collected using minimum dietary diversity questionnaire for women of reproductive age [9]. It entailed common consumed foods and frequency of foods intake. Mid upper arm circumference (MUAC) is a recognized test to determine pregnant woman’s nutritional condition [10] and was used to establish respondents nutrition status given that body mass index (BMI) is not recommend for use in pregnancy. Hemoglobin data levels was generated using a ham cue photometer by a competent medical laboratory technician at the pregnant adolescent households. Anaemia was defined as haemoglobin concentrations less than 120g/L after altitude correction in accordance with WHO recommendations, as none of the respondents were under the age of 12 [11].

#### Pre-testing of the questionnaire, validity and reliability

The tools’ administration time, content, and language were checked beforehand in Kajiado South’s Olgulului ward due to its similar cultural practices, such as early marriage, female genital mutilation, and socio-economic status. A sub-sample of 10% of the total sample was used for pretesting. Half of the sub-sample of 18 respondents were ANC users and with the other half being non-users. This did make it possible to modify repetition and unnecessary questions. A group of nutrition specialists and faculty from the department of Food, Nutrition, and Dietetics reviewed the questionnaire to elicit the required answers to assure validity. Validated questionnaires, calibrated blood analysers and anthropometric scales were used. The questionnaire was administered twice to 18 pregnant adolescents (about 10% of the sample group) to ensure reliability. After one week, the same sample was used in the follow-up test. An acceptable questionnaire did compare the findings and did have a reliability value of less than 0.7 [12]. A reliability coefficient of 0.84 determined using SPSS’s Cronbach’s alpha test, the questionnaire was deemed acceptable.

#### Questionnaire Administration

Data collection was done in Kajiado West, Ewauso and Keekonyoike wards. Four (4) diploma holders in nutrition and one (1) diploma holder in medical laboratory technician were hired and trained as research assistants. The training that took 3 days and it was done by the researcher using nutrition educational materials.

Upon arrival at the household (were the respondent was met) the research assistants did confirm the ages and pregnancy status to ensure eligibility. The questionnaire was administered to the participants by research assistants ensuring a thorough and consistent data collection process. On average, it took approximately 30 minutes to complete each questionnaire. To maintain confidentiality, the filled questionnaires were not left in the field but were securely transported for further analysis.

Data on dietary variety was collected using minimum dietary diversity questionnaire for women of reproductive age (MDD-W). It entailed common consumed foods and frequency of foods intake. The respondent did mention foods and the number of times she took in the previous 7 days. The data did also score adolescent pregnant foods consumed at least 5 out of the 10 food groups did achieve the recommended food intake.

The MUAC measurement was done on the less active midpoint of the upper arm while seated and relaxed. The recordings were done twice, later recorded to the nearest 0.1cm

A diploma holder in medical lab technician was used to collect the sample at homestead. Standard operating procedures (SOPs) for collecting blood samples were used [5]. A capillary blood sample was drawn using a sterile disposable lancet after cleaning the fingertip with an alcohol-soaked cotton swab. A micro cuvette was used to collect the second drop of blood once the first was wiped, then be placed in a portable Hemocue® battery-operated photometer (Hemocue® Hb 201+). In a minute the displayed value be recorded. Before each session, the photometer was calibrated. The blood was then placed in Ethylene Diamine Tetra Acetate coated vials stored in a cooler box temperature varying from 2 to 8 degrees Celsius safe from contamination before reaching the laboratory [5]. Training was given via role plays, fake interviews and demonstrations.

#### Focus Group Discussion (FGD) Guide

Focus group discussion (FGD) guide was used to collect qualitative data from the respondents. It was important to separate participants based on users and non-users to ensure the homogeneity of the group, which facilitated more effective discussions. Therefore, pregnant adolescent’s users and non-users were placed separate in their own FGD, separate from their parents, caregivers, or husbands. The study included randomly selected pregnant adolescents along with their parents/caregivers/husbands in groups of nine people each. They were later given FDG discussions guided by the researcher using guidelines and assisted by research assistant based on their attitudes, beliefs, and ideas, which provided more insights into the experiences of pregnant adolescent.

#### Key Informant Interview (KII) Guide

Key informant interview (KII) guides collected qualitative data from nurses in charge of each selected facility for ANC user and head teachers/ principals. Different guides were tailored for each group. Majorly the guide entailed nutrition knowledge, dietary practices, nutrition status, ANC attendance, IFAS consumption and adolescent transition in schools.

#### Data analysis plan

The study ensured the accuracy, completeness, consistency, reliability, and validity of the data, with any identified errors promptly resolved. Data was entered, cleaned, coded later labeled using the Statistical Package for Social Sciences (SPSS) software version 27, and analyzed at a significance threshold of 0.05. Data analysis did a comparison between the users and non-users of ANC. Data on income levels was influenced by location, occupation, education level and family size. Dietary practices, including meal frequency, dietary diversity and nutrient adequacy were investigated. The data is then analyzed using nutrisurvey software. Nutrient adequacy was assessed by comparing intake levels to Recommended Dietary Allowances (RDAs) as outlined by the [9] with specific cut-off points used to classify nutrient intake as adequate or inadequate. The RDA for adolescents include: calories 1400 to 2200 calories for ages 9-13 and 1800 to 2400 for ages14-18, carbohydrates 50-60% of the daily calories, protein 43-46g, fat 25-35% of daily calories, calcium 1300mg, iron 15mg, zinc 9 mg and vitamin A 700 µg, vitamin B1 1.0mg, vitamin B2 1.0mg, vitamin B6 1.2mg, vitamin B12 2.4, iodine 150 µg per day.

Dietary diversity was assessed using the Minimum Dietary Diversity for Women (MDD-W) indicator, categorizing intake into 10 food groups namely grains products/starchy foods, legumes and pulses, dairy products, eggs, flesh, green leafy vegetables, nuts and seeds, yellow orange fruits and vegetables, other vegetables and other fruits with a cut-off point of 5 or more food groups out of 10 to indicate adequate dietary diversity, following [9]. Data on Iron and Folic Acid Supplementation (IFAS) consumption were measured by the frequency of intake, specifically assessing whether participants consumed supplements at least three times per week, adhering to [11]. ANC attendance and IFAS intake was positively addressed.

Nutrition status was evaluated using Mid-Upper Arm Circumference (MUAC) < 22cm did showed underweight and >22cm was normal nutrition status [13].The filled questionnaires were not left in the field to maintain confidentiality but were securely transported for further analysis. To identify correlations between variables, the Pearson product moment correlation test was employed, and associations among variables were determined using the Chi-square test. Logistical regression did predict outcomes while controlling for confounding variables. The data was evaluated at the 0.05 significance level. FDG and KII were transcript from the recordings, notes were organized into the same subjects, and a summary was prepared for each [14]

### Data analysis plan

#### Sampling Procedure

The sampling was conducted using multi-stage random sampling. Kajiado West sub-county region was purposively selected due to high levels of poverty and frequent food supply shocks, ensuring the study focuses on areas significantly impacted by these issues. Random sampling was done in the 5 wards namely Ewauso, Iloodokilanik, Keekonyoike, Magadi and Mosiro where Ewauso and Keekonyoike were chosen.

Data was collected from 19 health institutions of the randomized sample of 38 health facilities from Ewauso (24) and Keekonyoike (14) wards which aided in getting records of 98 ANC pregnant adolescent users in both wards. Records of ANC pregnant adolescent users were used. A total of 210 pregnant adolescent ANC users were recovered from health facility records of which they were randomly sampled on every 2^nd^ household. The 38 health institutions linked respective schools where ANC non-users were identified. Ewauso having 22 schools and keekonyoike 21 schools.

Community Health Promotors (CHP) did also aided in identifying the ANC non-users in the wards. CHPs and records from schools identified 150 households with ANC non-users. In Keekonyoike ward, a total of 60 households were identified and 90 households in Ewauso ward. Simple random sampling was used to choose from 150 household with pregnant teens in the two wards. The first household was chosen using a table of random numbers and each n^th^ number (every 2^nd^) household was drawn to achieve 98 ANC non-users. Hemoglobin (Hb) levels were determined by analysis of blood samples taken from all participants by a competent medical lab technician.

## Results

### 4.0. Introduction

Out of the calculated sample size of 196, the study had 189 responses, resulting in a response rate of 96.4%. Of the 189 respondents whose responses were utilized for the study, a majority (68.8%; n=130) indicated that they used ANC services during pregnancy, while about a third (31.2%; n=59) reported not using ANC services at any point during their pregnancy

### Demographics and Socio-economic of Pregnant Adolescents in Kajiado

#### Demographics Characteristics of the Pregnant Adolescents by ANC Utilization

Less than a half (39.7%) lived with their parents having users (30.8%) and non-users 59.3%. More than half (53.4%) did live their spouse having (62.3%) users and (33.9%) non-users. Minority (1.6%) were single having (1.5%) users and 1.7% non-users. Minority (5.3%) others having (5.4%) users and (5.1%) non-users. The difference between the two groups was statistically significant, with a (p-value of 0.554) (Table 1).

**Table 1.** Distribution of Respondent Users and Non-users of ANC Clinics by Demographic Characteristics.

| Demographic characteristics | Total (N=189) | | Users of ANC (N=130) | | Non-Users of ANC (N=59) | | $\chi^2$ (P-value) |
| --- | --- | --- | --- | --- | --- | --- | --- |
|  | N | % | n | % | n | % |  |
| **Age (years)** |  |  |  |  |  |  |  |
| 0≤14 | 4 | 2.1 | 3 | 2.3 | 1 | 1.7 | 0.415 |
| 15-16 | 69 | 36.5 | 48 | 36.9 | 20 | 33.9 |  |
| 17-19 | 116 | 61.4 | 79 | 60.8 | 38 | 64.4 |  |
| **Currently living with** |  |  |  |  |  |  |  |
| Parent | 75 | 39.7 | 40 | 30.8 | 35 | 59.3 | 0.554 |
| Spouse | 101 | 53.4 | 81 | 62.3 | 20 | 33.9 |  |
| Single | 3 | 1.6 | 2 | 1.5 | 1 | 1.7 |  |
| Others | 10 | 5.3 | 7 | 5.4 | 3 | 5.1 |  |

\* Others currently living with included; siblings, extended family
\*Other religion include; not attending any religion

#### The Socio-economic Status of Households of Pregnant Adolescents by ANC Utilization

More than half (57.7%) of the households earned less than KSh. 2,000 per month having (60.8%) ANC users (54.2%) non-users. Slightly more than half (50.8%) of the households earned KSh. 2,001–5,000 per month having (11.5%) ANC users and (28.8%) non-users. Minority (10.6%) of the pregnant adolescent earned between KSh. 5,001–10,000 having (13.1%) ANC users and (5.1%) non-users. A smaller number (10.0%) earned KSh. 10,001–20,000 having (11.5%) ANC users and (3.4%) non-users. A small number (4.8%) earned above KSh. 20,001 having (3.1%) users and (8.5%) non-users. Statistical significant was indicated between the two groups (p-value 0.008).

More than two thirds (63.5%) had mud-walled houses having, (70.8%) ANC users and (47.4%) non-users. Nearly quarter (21.2%) walls were made of wood, having (13.8%) ANC users and (37.3%) non-users. Minority (15.3%) walls were stoned/cemented having (15.4%) ANC users and (15.3%) non-users. The differences between the two groups was statistically significant having a (p-value of 0.810) (Table 2).

**Table 2.** Distribution of Respondent Users and Non-users of ANC Clinics by Socio-economic Characteristics.

| Socio-economic characteristics | Total (N=189) | | Users of ANC (N=130) | | Non-Users of ANC (N=59) | | $\chi^2$ (P-value) |
| --- | --- | --- | --- | --- | --- | --- | --- |
|  | n | % | n | % | n | % |  |
| **Household income** |  |  |  |  |  |  |  |
| Less than 2,000 | 109 | 57.7 | 79 | 60.8 | 32 | 54.2 | **0.008** |
| 2,001 - 5,000 | 32 | 16.9 | 15 | 11.5 | 17 | 28.8 |  |
| 5,001 - 10,000 | 20 | 10.6 | 17 | 13.1 | 3 | 5.1 |  |
| 10,001 - 20,000 | 19 | 10.0 | 15 | 11.5 | 2 | 3.4 |  |
| 20,001 and above | 9 | 4.8 | 4 | 3.1 | 5 | 8.5 |  |
| **Main cooking material** |  |  |  |  |  |  |  |
| Charcoal | 92 | 48.8 | 57 | 43.8 | 35 | 59.3 | 0.090 |
| Wood & Shrub | 91 | 48.1 | 68 | 52.3 | 23 | 39.0 |  |
| Charcoal, wood & shrubs | 5 | 2.6 | 4 | 3.1 | 1 | 1.7 |  |
| Biogas | 1 | 0.5 | 1 | 0.8 | 0 | 0.0 |  |
| **Material on the Floor of the House** |  |  |  |  |  |  |  |
| Cement/Ceramic | 122 | 64.6 | 87 | 66.9 | 35 | 59.3 | **0.003** |
| Earth/Sand | 17 | 8.9 | 2 | 1.6 | 15 | 25.4 |  |
| Wood | 50 | 26.5 | 41 | 31.5 | 9 | 15.3 |  |
| **Exterior Walls** |  |  |  |  |  |  |  |
| Mud | 120 | 63.5 | 92 | 70.8 | 28 | 47.4 | 0.810 |
| Wood | 40 | 21.2 | 18 | 13.8 | 22 | 37.3 |  |
| Stone/Cement | 29 | 15.3 | 20 | 15.4 | 9 | 15.3 |  |

#### Dietary Practices of Pregnant Adolescents by ANC Utilization

Table 3 presents dietary practices of pregnant adolescent by ANC utilization. Majority (98.9%) of the respondent consumed grains products/starchy foods having (69.0%) ANC users and (31.0%) non-users showing no significant difference (p-value 0.564) between the two groups. Over a third (93.7%) of the respondent consumed legumes and pulses having (68.9%) ANC users and (31.1%) non-users showing here was no significant difference (p-value 0.87) between the two groups.

**Table 3.** Distribution of Respondent Adolescent Users and Non-users of ANC by Food Group Consumption.

| **Food groups** | **Total**<br>**n (%)** | **Users**<br>**n (%)** | **Non-Users**<br>**n (%)** | **$\chi^2$**<br>**(P-value)** |
| --- | --- | --- | --- | --- |
| Grain Products/Starchy foods | 187(98.9) | 129(69.0) | 58(31.0) | 0.564 |
| Legumes and Pulses | 177(93.7) | 122(68.9) | 55(31.1) | 0.87 |
| Nuts and seeds | 13(6.9) | 10(76.9) | 3(23.1) | 0.512 |
| Dairy | 189(100.0) | 130(68.8) | 59(31.2) | **0.00** |
| Meat, poultry and fish | 126(66.7) | 87(69.0) | 39(31.0) | 0.912 |
| Eggs | 83(43.9) | 58(69.9) | 25(30.1) | 0.773 |
| Dark green leafy vegetables | 183(96.8) | 124(67.8) | 59(32.2) | 0.094 |
| Vitamin A-rich fruits and vegetables | 151(79.9) | 106(70.2) | 45(29.8) | 0.402 |
| Other vegetables | 185(97.9) | 127(68.6) | 58(31.4) | 0.786 |
| Other fruits | 109(57.7) | 66(60.6) | 43(39.4) | **0.009** |
| **Equal or greater than 5 food groups (N=189)** | **(N=189)** | **(N=130)** | **(N=59)** |  |
| Pregnant Adolescent | 186(98.4) | 127(68.3) | 59(31.7) | 0.24 |

[10] Minimum dietary diversity for women. In *Minimum dietary diversity for women* (Vol. 1). FAO. https://doi.org/10.4060/cb3434en

Minority (6.9%) of the respondent consumed nuts and seeds having (76.9%) ANC users and (23.1%) non-users showing no significant difference (p-value 0.512) between the two groups. Majority (100.0%) of the respondent consumed dairy products having (68.8%) ANC users and (31.2%) non-users, showing extremely unlikely under the assumption (p-value 0.00) between the two groups.

More than three quarter (79.9%) of the respondent consumed yellow orange fruit and vegetables having (70.2%) ANC users and (29.8%) non-users. There was no significant difference (p-value 0.094) between the two groups. Majority (79.9%) of the respondent consumed other vegetables having (68.6%) ANC users and (31.4%) non-users showing no significant difference (p-value 0.786) between the two groups. More than half (57.7%) of the respondent consumed fruits having (60.6%) ANC users and (39.4%) non-users. There was a significant difference (p-value 0.009) between the two groups.

Majority (98.4%) of the respondent consumed equal or greater than 5 food groups other having (68.3%) ANC users and (31.7%) non-users. There was no significant difference (p-value 0.24) between the two groups (Table 3).

#### Energy Proteins and Fats of Pregnant Adolescents by ANC Utilization

Table 2 presents nutrient intake analyses of pregnant adolescent. The proportion meeting energy (1086.8 ± 90.57 Kcal) (RDA 29.0%) having (1105.72 ± 95.43 Kcal) (RDA 29.6%) users and (1070.12 ± 8.36 Kcal) (RDA 28.4%) non-users was recommended daily amount (RDA) showing a significant differences (p= 0.01).

Protein intake was (47.46 ± 3.96 g) (RDA 31.5%) having (49.16 ± 0.12 g) (RDA 32.5%) users and (45.76 ± 3.84 g) (30.5% RDA) non-users showing a significant differences (p= 0.023). Fat intake was (40.76 ± 3.40 g) (RDA 29.3%) having (42.32± 3.57 g) (RDA 30.1%) users and (39.20 ± 3.24 g) (28.5% RDA) non-users. There was a significant between the two groups (p-value 0.024). Vitamin A intake was (620.22 ± 50.19µg) (RDA 39.9%) having (620.22 ± 51.73µg) (RDA 40.9%) users and (584.22 ± 48.65µg) (RDA 38.9%) non-users thus showing significant between the two groups (p-value 0.019). Vitamin B1 intake was (1.12± 0.093mg/day) (RDA 40.2%) having (1.15± 0.10mg/day) (RDA 40.2%) (RDA 41.2%) and non-users (1.09 ± 0.091mg/day) (RDA 39.2%) thus showing significant difference (p-value 0.017).

Iron intake was (7.61± 0.63mg/day) (RDA 39.3%) having (7.91± 0.66mg/day) (RDA 40.3%) and non-users (7.31 ± 0.060mg/day) (RDA 38.3%) thus showing significant difference (p-value 0.025). Iodine intake was (145± 12µg) (RDA 34.7%) having (150 ± 12.5µg) (RDA 35.2%) users and (140 ± 11.5µg) (RDA 34.2%) non-users, having a statistical significant between the two groups (p-value 0.022). Zinc intake was (7.9± 0.65mg/day) (RDA 30.8%) having (8.1± 0.68mg/day) (RDA 31.3%) and non-users (7.8 ± 0.063mg/day) (RDA 30.2%) thus showing significant difference (p-value 0.012) (Table 4).

**Table 4.**
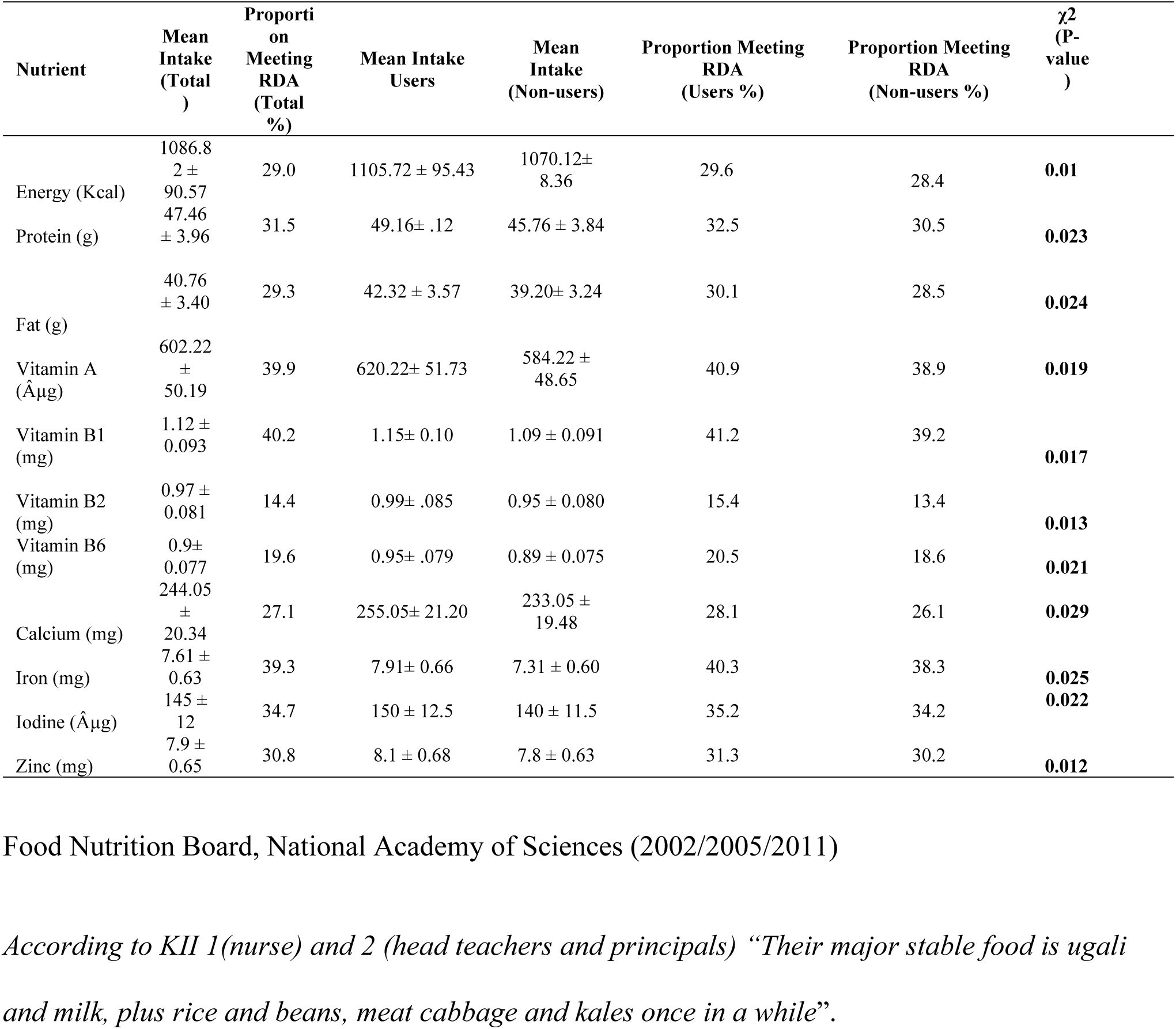
Distribution of Respondent Adolescent Users and Non-users of ANC by Energy and Nutrient Intake.

#### Factors Associating Dietary Practices of Pregnant Adolescents by ANC Utilization

Table 5 presents factors associating dietary practices among pregnant adolescents.

**Table 5.**
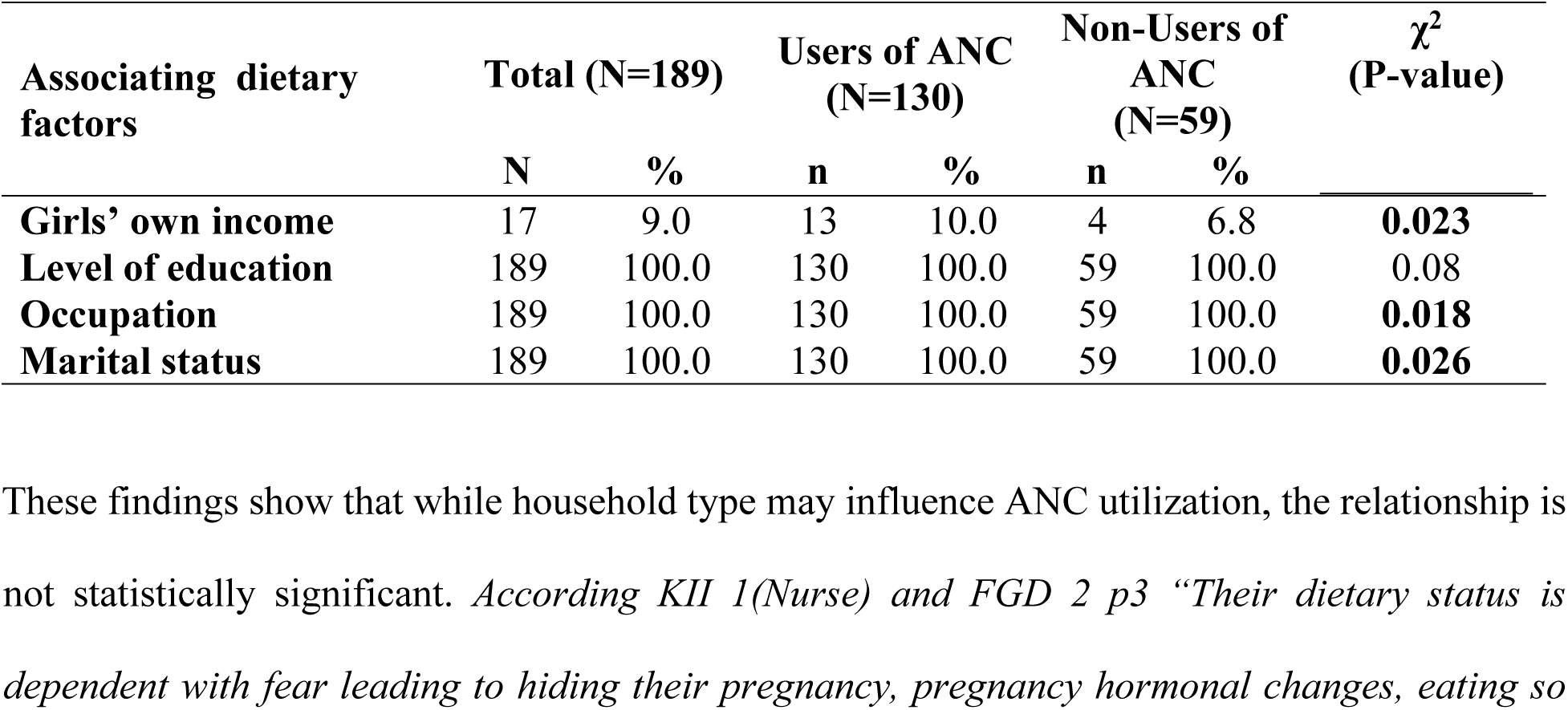
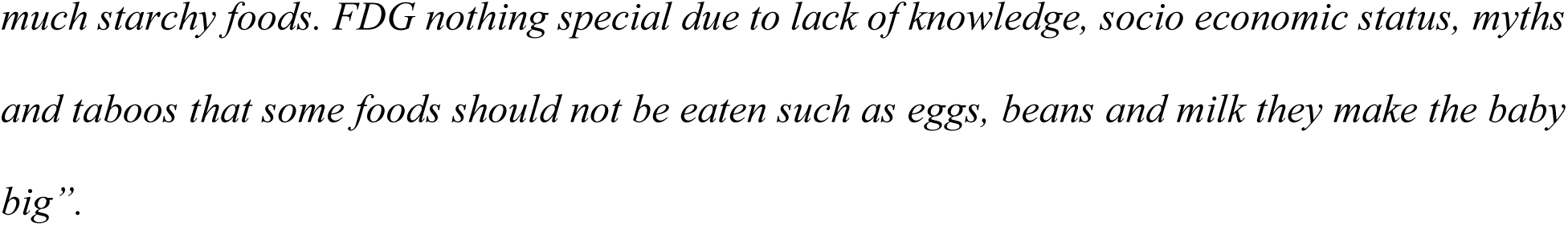
Distribution of Respondent Adolescent Users and Non-users of ANC by Associating Dietary Factors.

Minority (9.0%) of pregnant adolescent association with their own income (10.0%) users and (6.8%) non-users. There was significant difference between the two groups (p-value 0.023). Majority (100.0%) of pregnant adolescent association with level of education had their own income (100.0%) users and (100.0%) non-users. There was no significant difference between the two groups (p-value 0.08).

Majority (100.0%) of pregnant adolescent association with occupation (100.0%) users and (100.0%) non-users. There was significant difference between the two groups (p-value 0.018). Majority (100.0%) of pregnant adolescent association with marital status (100.0%) users and (100.0%) non-users. There was no significant difference between the two groups (p-value 0.026).

### Nutrition Status of Pregnant Adolescents

#### MUAC Status of Pregnant Adolescents by ANC Utilization

Table 6 presents adolescent nutritional status measured using MUAC since BMI is not used in pregnancy. MUAC cut points with malnutrition level are WHO. Nearly a quarter (31.2%) of respondents having (MUAC < 22 cm) indicated potential malnutrition or nutritional risk at (30.8%) ANC users at (30.8%) non-users (32.2%). Majority (68.8%) of pregnant adolescents had normal nutritional status (MUAC > 22 cm) of all participants (69.2%) ANC users (67.8%) non-users had normal nutrition status showing no significant difference (p= 0.534) (Table 6).

**Table 6.** Distribution of Respondent Adolescent Users and Non-users of ANC by MUAC Nutrition Status.

| **MUAC Status (cm)** | **Total**<br>**(N=189)**<br>**n (%)** | **Users**<br>**(N=130)**<br>**n (%)** | **Non-Users**<br>**(N=59)**<br>**n (%)** | **$\chi^2$**<br>**(P-value)** |
| --- | --- | --- | --- | --- |
| Underweight (MUAC < 22 cm) | 59 (31.2) | 40 (30.8) | 19 (32.2) | 0.534 |
| Normal Nutritional Status (MUAC > 22 cm) | 130 (68.8) | 90 (69.2) | 40 (67.8) |  |

#### Hemoglobin Status of Pregnant Adolescents by ANC Utilization

Table 7 shows nutritional status of the pregnant adolescents, based on their hemoglobin levels. Less than half (42%) of the pregnant adolescent had low (<11.5g/dl) having (30.0%) ANC users and (12.0%) non-users. Majority of respondent had normal (11.5-13.5g/dl) having (63.0%) ANC users and (39.0%) non-users.

**Table 7.**
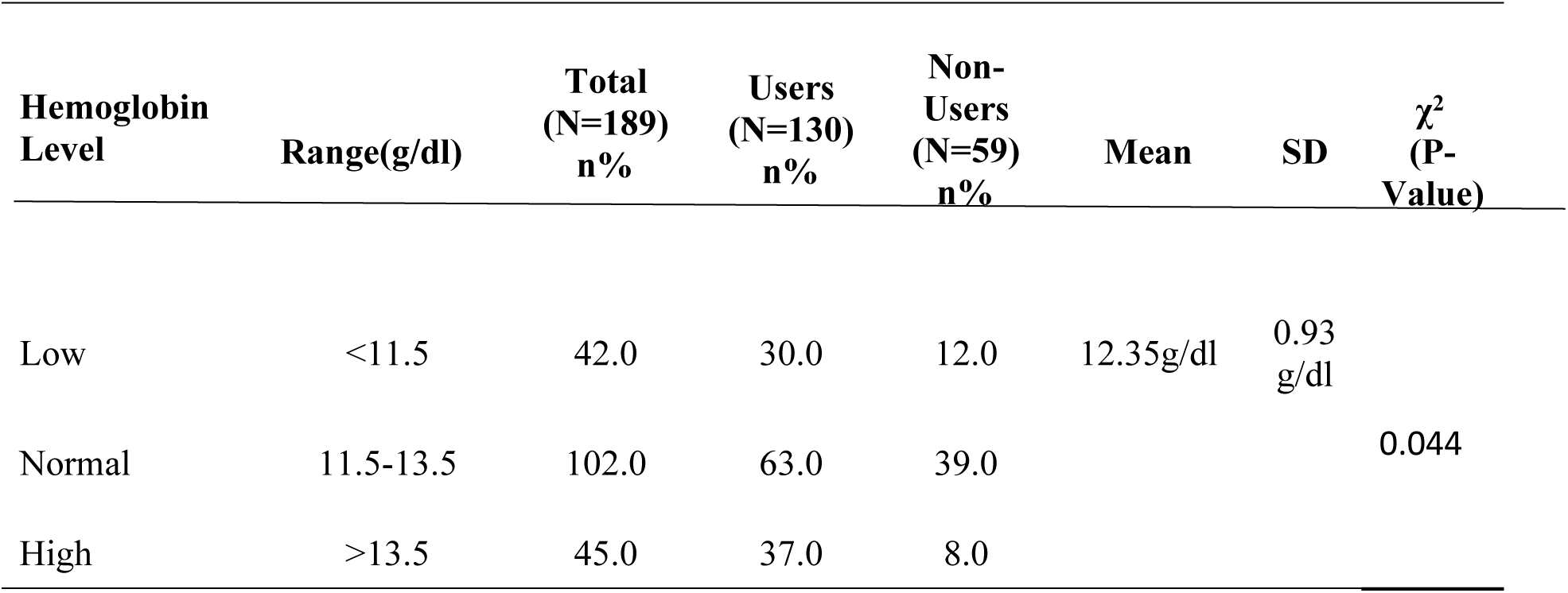
Distribution of Respondent Adolescent Users and Non-users of ANC by Hemoglobin Level.

Less than half (45.0%) had high (<13.5g/dl) having (37.0%) ANC users and (8.0%) non-users with significant difference between the two groups (p-value 0.044) (Table 7).

### Factors Affecting Attendance of ANC and Consumption of IFAS among Pregnant Adolescent

#### Factors Affecting ANC Attendance among of Pregnant Adolescent by ANC Utilization

Table 8 presents factors affecting attendance of ANC and consumption of IFAS.

**Table 8.**
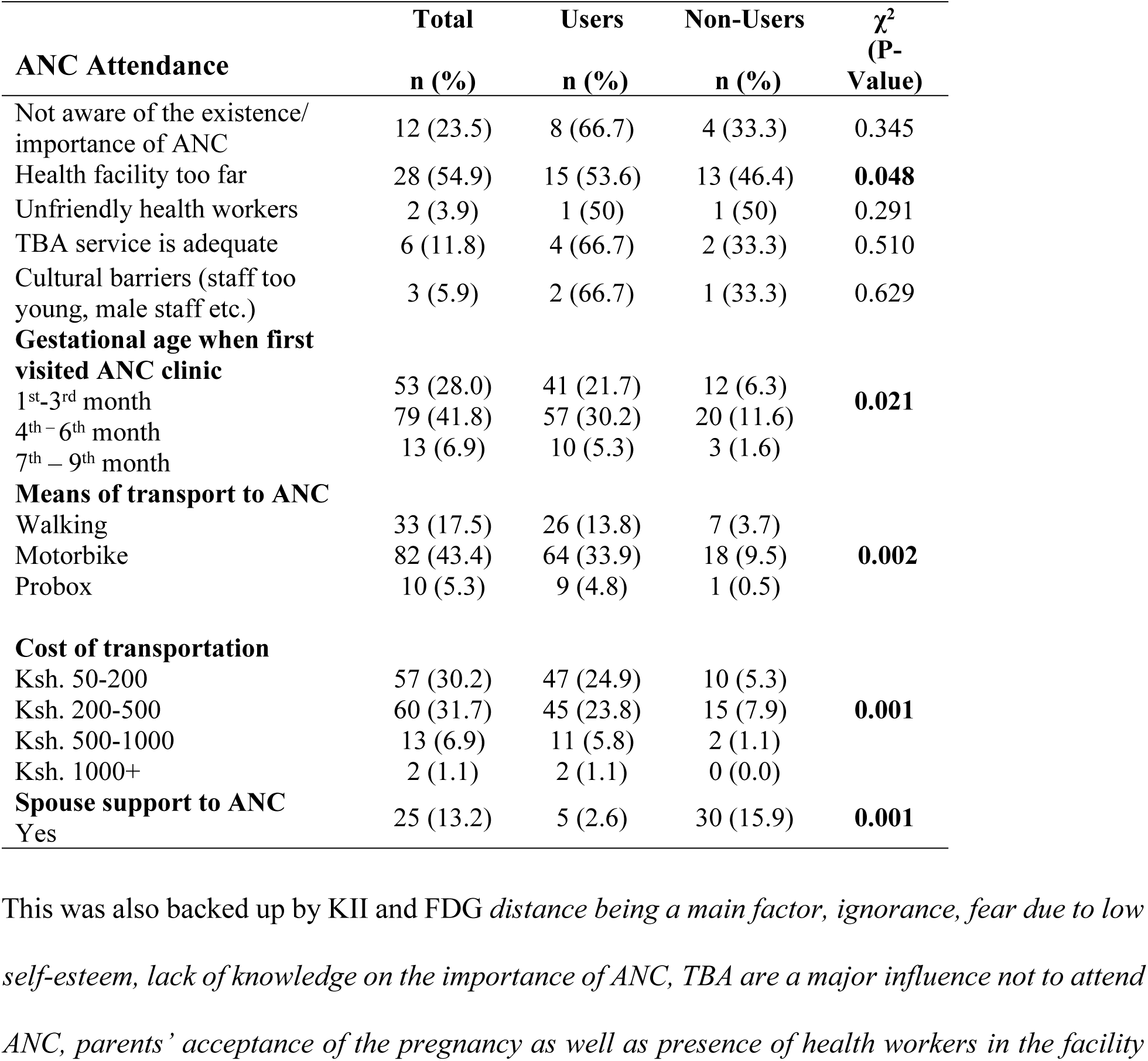
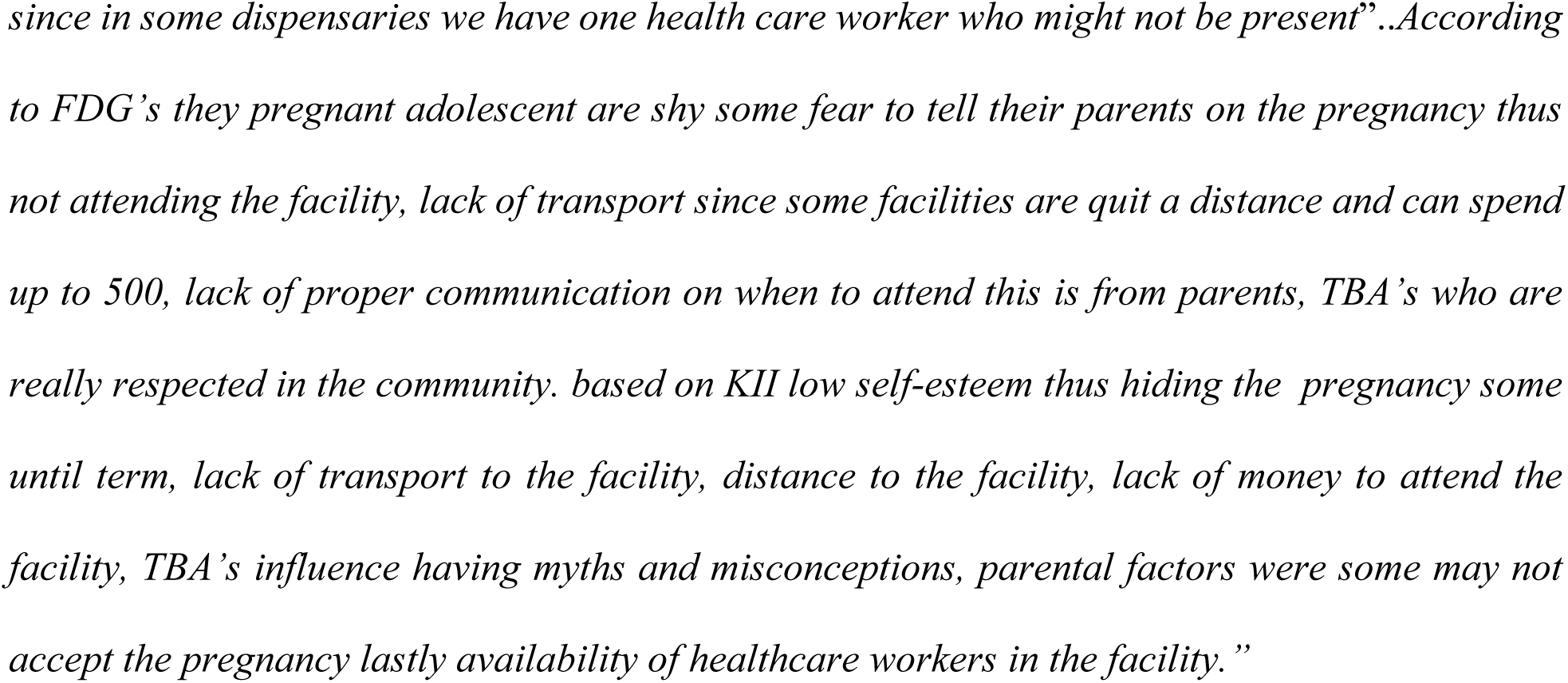
Distribution of Respondent Adolescent Users and Non-users of ANC-by ANC Attendance.

Less than a quarter (23.5%) of the pregnant adolescent were not aware of the existence/importance of ANC having (66.7%) ANC users and (33.3%) non-users showing no significant difference between the two group (p-value 0.048).

More than a half (54.9%) of the pregnant adolescent claimed that the health facilities were too far having (53.6%) ANC users and (46.4%) non-users showing there was significant difference between the two group (p-value 0.048).

Minority (3.9%) of the pregnant adolescent claimed ANC attendance was affected by unfriendly staffs (50.0%) ANC users and (50.0%) non-users. There was no significant difference between the two groups (p-value 0.291). A small number (11.8%) relied on TBA having (66.7%) ANC users and (33.3%) non-users showing there was no significant difference between the two group (p-value 0.510).

A small number (5.9%) of claimed cultural barriers affected such as male staffs, staff too young having (66.7%) ANC users and (33.3%) non-users showing there was no significant difference between the two group (p-value 0.629). More than a quarter (28.0%) did visit ANC gestational age 1^st^-3^rd^ month having (21.7%) ANC users and (6.3%) non-users. Less than a half (41.8%) 4^th^-6^th^ month having (30.2%) and (11.6%). Minority (6.9%) 7^th^-9^th^ months having (5.3%) and (1.6%) non-users showing a significant difference between the two group (p-value 0.021).

A small number (17.5%) of pregnant adolescent used walking as means of transport to ANC having (13.8%) and (3.7%) non-users. Less than half (43.4%) of pregnant adolescent used motorbike having (33.9%) and (9.5%) non-users. Minority (5.3%) used probox having (4.8%) and (0.5%) non-users. These did show a significant difference between the two groups (p-value 0.002).

More than quarter (30.2%) of pregnant adolescent’s monthly income was Ksh 50-200 having (24.9%) ANC users and (5.3%) non-users. More than quarter (31.7%) of pregnant adolescent’s monthly income was Ksh 200-500 having (23.8%) ANC users and (7.9%) non-users. These did show a significant difference between the two groups (p-value 0.001).

#### Factors Affecting Consumption of IFAS Pregnant Adolescent by ANC Utilization

Table 9 presents factors affecting consumption of Iron Folic Acid Supplements (IFAS) for 7 days in a week. Minority (1.1%) did not have enough IFAS tablets having (0.5%) ANC users and (0.5%) non-users. A small number (2.6%) didn’t know how take IFAS, having (2.1%) ANC users and (0.5%) non-users. Less than a quarter (11.6%) having (6.3%) ANC users and (5.3%) non-users. A small number (1.1%) claimed to forget and having side effects (1.1%) ANC users and (0.0%) non-users, for those who has side effects (3.7%) having (3.2%) ANC users and (0.5%) non-users. There was a statistical significant difference between the two groups (p-value 0.041).

**Table 9.** Distribution of Respondent Adolescent Users and Non-users Factors Affecting IFAS Consumption.

| Factors influencing consumption | Total<br>n (%) | Users<br>n (%) | Non-Users<br>n (%) | $\chi^2$<br>(P-value) |
| --- | --- | --- | --- | --- |
| Did not have enough IFAS tablets | 2 (1.1) | 1 (0.5) | 1 (0.5) | **0.041** |
| Didn't know how long to take the tablets | 5 (2.6) | 4 (2.1) | 1 (0.5) |  |
| Forgot | 22 (11.6) | 12 (6.3) | 10 (5.3) |  |
| Forgot and Side effects | 2 (1.1) | 2 (1.1) | 0 (0.0) |  |
| Side effects | 7 (3.7) | 6 (3.2) | 1 (0.5) |  |

*According to KII's and FDG's done the side effects thus not taking, importance of taking IFAS, forgetting to take IFAS, lack of transport to attend the next visit to pick IFAS thus defaulting, still have misconceptions from men that it's a form of family planning, as per parity they tend to ignore since if they get a healthy baby they don't see they need to take in the next pregnancy, sometime IFAS is not available in the facility."*

#### Relationship between Dietary Practices and Nutrition Status of Pregnant Adolescents

The analysis showed statistically significant associations, between marital status, level of education, age and household income (p-values < 0.001).When examining the dietary practice, there is statistical significant correlation with age (p-value 0.004), income (p-value 0.005), and the dietary diversity score itself (p-value 0.014).

The results indicate a significant relationship between socio-economic characteristics, which include household income, marital status, and education, and the nutrition status of pregnant adolescents. The analysis revealed that both household income and marital status have a significant association with the nutrition status of the adolescents (p-value < 0.001).

Lastly, the analysis of IFAS consumption and nutrition knowledge revealed significant relationship (p-value < 0.001). Pregnant adolescents who had higher levels of nutrition knowledge were more likely to regularly consume IFAS, which is critical for preventing anemia and ensuring proper fetal development. The analysis also showed that adolescents who understood the benefits of IFAS were more compliant with their intake, compared to those who lacked knowledge (Table 4.15).

**Table 10.** Relationship between Nutrition Knowledge and Dietary Practices.

| Relationship between variables | Variable | $\chi^2$<br>(P-value) |
| --- | --- | --- |
| Relationship between knowledge score and other variables | Marital status | **p= <0.001** |
|  | Level of education | **p= <0.001** |
|  | Age | **p= 0.081** |
|  | Household income | **p= <0.001** |
|  | Age | **p= 0.004** |
| Relationship between dietary practice score and other variables | Nutrition status | p=0.302 |
|  | Hb levels | **p = 0.021** |
|  | Income | **p= 0.005** |
|  | Dietary diversity score | **p= 0.014** |
| Household, income, marital status, Education and nutrition status |  | **p=<0.001** |
| IFAS consumption and nutrition Knowledge |  | **p=<0.001** |

#### Relationship between Nutrient Intake and Nutrition Status

The correlation analysis between nutrient intake and nutrition status reveals varying relationships. Vitamins and Mid-Upper Arm Circumference (MUAC) (p-value 0.050), suggesting there was no significant difference. However, there was a significant difference between carbohydrate intake and MUAC (p-value 0.023), indicating that higher carbohydrate intake is associated with higher MUAC. Proteins and MUAC showed a significant difference (p-value 0.018) also suggesting that higher proteins intake is associated with higher MUAC. This finding suggests that carbohydrates and proteins may play a significant role in influencing MUAC compared to vitamins (Table 11).

**Table 11.**
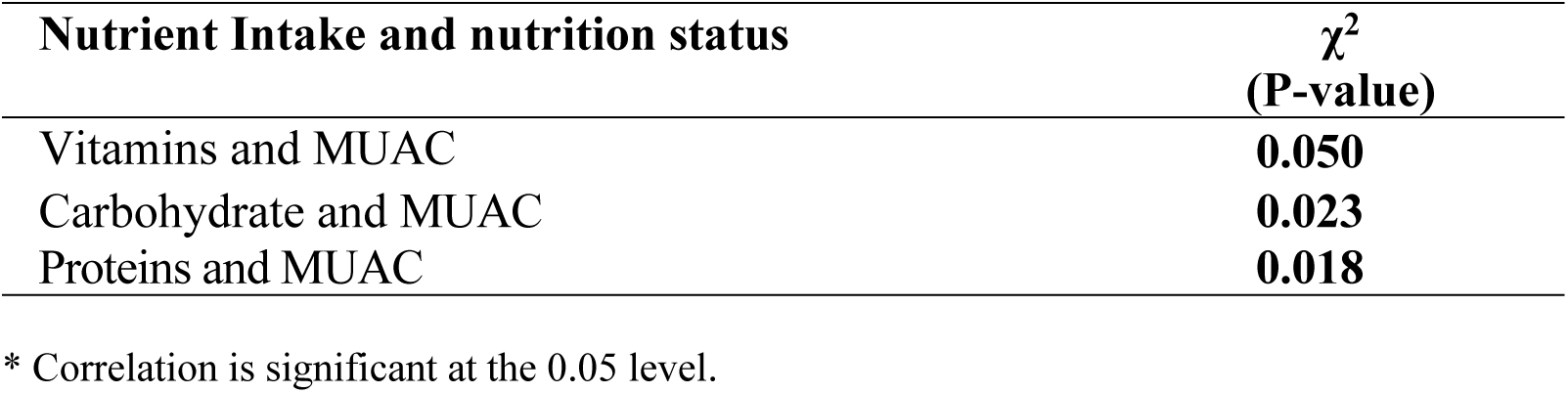
Nutrient Intake and Nutrition Status.

#### Relationship between Dietary Practices and Nutrition Status of Pregnant Adolescents

Dietary practice analysis showed statistical significant correlation with age (p-value 0.004), income (p-value 0.005), and the dietary diversity score itself (p-value 0.014).

The results indicate a significant relationship between socio-economic characteristics, which include household income, marital status and the nutrition status of pregnant adolescents. The analysis revealed that both household income and marital status has a statistical significant association with the nutrition status of adolescents (p-value < 0.001) (Table 12).

**Table 12.**
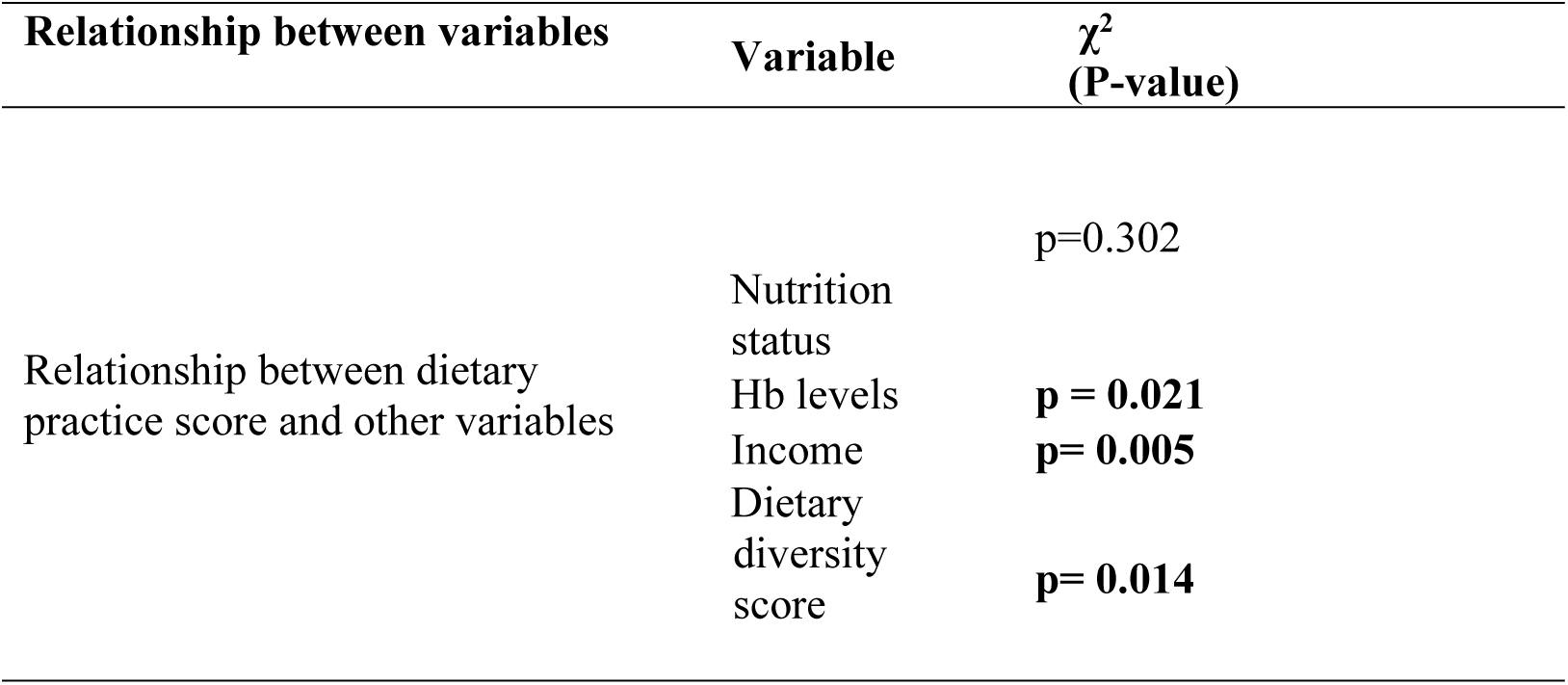
Relationship between Dietary Practices.

| Relationship between variables | Variable | $\chi^2$<br>(P-value) |
| --- | --- | --- |
| Relationship between dietary practice score and other variables | Nutrition status | p=0.302 |
|  | Hb levels | **p = 0.021** |
|  | Income | **p= 0.005** |
|  | Dietary diversity score | **p= 0.014** |

## Discussion

### Demographic, and Socio-Economic Characteristics of Pregnant Adolescents Demographic Characteristics

Adolescence is a crucial time of remarkable changes, including fast maturation of the physical, psychological, sexual, and cognitive domains. This study did concur with [15] that adolescents also have greater nutritional needs during this time than at any other point in their lives. Adolescent females who are expecting are especially vulnerable because they have additional dietary needs to sustain a growing fetus on top of the demands of normal growth and development. The study deviates from [16] findings, which suggest that early marriage is associated with a lower nutritional status in early pregnancy (≤15 years of age) as opposed to late pregnancy. Further, as established by the study findings, the majority of respondents (39.8%) were housewives.

The study’s findings also suggest that housing conditions may play a role in ANC utilization. Respondent (64.6%) had cement or ceramic floors in their homes, which were significantly more common among ANC users (66.9%) compared to non-users (59.3%). This correlation between housing quality and ANC service use might reflect broader socio-economic disparities that affect overall health, as higher-quality housing is often linked with improved access to healthcare [17].

Additionally, main cooking materials varied between ANC users and non-users, with (48.8%) of respondents using charcoal, and (48.1%) using wood and shrub. While no statistically significant difference was found in cooking materials (p-value = 0.090), this aspect of household infrastructure can influence health outcomes, as different cooking methods might impact maternal nutrition, as discussed by [18].

The study found that the majority of respondents (68.3%) of adolescents who utilized antenatal care (ANC) services consumed foods and beverages from five or more food groups on the previous day, compared to only 31.7% of women who did not use ANC services. The dietary pattern analysis approach provides a more comprehensive understanding of dietary habits and helps explain how socioeconomic factors influence adherence to these patterns, which include combinations of commonly consumed foods.

Iron deficiency anemia could be caused by various factors, which include poor dietary intake of iron-rich foods, unavailability rich iron foods, household, food and nutrition insecurity, low nutrition knowledge and attitudes, high demand for iron during pregnancy, infestations of worms and infections such as malaria [19]. Furthermore, most pregnant women in developing nation already have low levels of iron in their blood, which is exacerbated by the developing baby’s increased requirement for iron. It is alarming that pregnant women’s diet of iron-rich animal foods may raise their risk of iron deficiency anemia because haeme iron, which is found in meat, is more accessible than non-haeme iron, which is found in plants. This study concurs [20] limited consumption of these iron-rich meals could be attributed to their high cost, as most pregnant women who reported difficulty preparing iron-rich foods listed cost as a barrier. Farm animals are retained in disadvantaged rural areas like the one under study for economic reasons rather than for consumption.

The study [17] describes poor dietary patterns among pregnant teens. This is due to a small number of meals ingested each day, a lack of diversification of the diet, inadequate intake of important nutrients required for optimum nutrition during pregnancy. Poor dietary understanding has contributed to the high prevalence of underweight. Another noteworthy conclusion from the current study [21] was the restricted consumption of animal-based foods despite the increasing need for high equality, sufficient protein during pregnancy. The relationship between maternal dairy product consumption and fetal development concludes that lower cost animal source foods such as milk (which is consumed regularly) may have more positive impact in maternal nutrition and fetal growth than meats, poultry, and fish products, which are expensive and consumed only occasionally.

The study’s energy RDA was 29.0 percent. The current study found an overreliance on carbohydrates for daily energy, most likely due to a preference for maize-based foods such as rice and mandazi. The nutritional needs of adolescents are considerable and are at risk of malnutrition (underweight, overweight, micronutrient deficiencies) [22]. Iron deficiency disproportionately affects pregnant adolescents with more than 30% living in countries with poorer social development index (SDI) ratings. In these countries, vitamin A deficiency affects 20% of girls aged 10-14 and 18% of girls aged 15-19, while iodine deficiency affects 3% and 5% of younger and older girls, respectively. A comparable study in China found that pregnant women had low dietary intakes of both calories, vitamin A, iron, and zinc. According to [23] how many meals consumed is related to diversity of diet, energy intake, and nutrient content. Dietary diversity also influenced vitamin and mineral consumption. Consuming fruits and vegetables has been shown to boost vitamins and minerals intake, including vitamin A, iron, and zinc.

This study [17] reveals unhealthy eating habits among pregnant teenagers. This is due to the limited meals consumed daily, inadequate dietary diversity, and not frequently consuming key nutrients required for optimal nutrition during pregnancy. However, the current study’s dietary data was assessed to be equal, as other research have also found lower food intake when compared to the RDI and RDA for pregnancy.[22] reported intake of less than 50% of the RDA. According to [24] discovered that dietary diversity was low among pregnant women and was substantially correlated with education and socioeconomic status.

Women from the lower-middle and low classes had medium diet variety scores when compared to those from the middle class. The study by [24] demonstrated a significant (p < 0.01) association between mother calorie and protein intake during pregnancy and infant birth weight. Infant weight climbed steadily as the mother’s calorie and protein consumption increased. Maternal micronutrient consumption (riboflavin, niacin, B6, and zinc) during pregnancy was found to have a significant relationship with newborn anthropometry. Adolescent girls are particularly prone to iron deficiency anemia due to their rapid growth during adolescence [24], which corresponds with the onset of menstruation. Pregnant teenagers are more vulnerable because of the increased requirement for iron during pregnancy to expand maternal tissues and promote fetal growth.

These findings imply that pregnant teenagers are often unaware of the need of diet for a healthy pregnancy; they also lack comprehension of the precise links between maternal diet and fetal development, as many did not change their diet after becoming pregnant [25] Cravings, food availability, taste preferences, and financial considerations were among the most frequently mentioned factors influencing dietary consumption. While pregnant teens relied on others for food, the majority reported having the most control over what they ate.

Overall, this group of pregnant teens defined healthy eating as limiting snacking, eating regularly, and adding fruits and vegetables in their meals. Despite this information, prior research from this cohort suggested that most teens did not achieve the projected average requirements for prenatal nutrients such as vitamins E and D, calcium, magnesium, and iron, while also surpassing sugar recommendations, particularly for added sugars. [25] findings support prior research indicating pregnant teenagers frequently skip meals and graze on energy-dense convenience items that are deficient in micronutrients..

According to the study, low socioeconomic status was (74.6%), primary level (52.9%), and cultural views had the greatest influence (68.8%). Health workers were (52.9%) effective in disseminating health education, which altered eating practices. Evidence suggests that a variety of factors influence pregnant women’s food preferences. Culture, as previously said, influences people’s dietary choices, and certain societies prohibit the consumption of particular foods, such as rabbit meat. Food taboos may influence certain women’s eating patterns, leading them to shun certain foods. Food taboos have the potential to negatively affect women’s nutritional status [26].

In China, a research of pregnant women found that the study participants were more likely to follow traditional diet advice such as avoiding particular items like rabbit meat [27]. A research of pregnant women in Khartoum found that participants avoided consuming certain meals due to dietary taboos, personal and community reasons [28]. Reasons are presented to support the taboos and food restrictions. In Tanzania, a study of pregnant Maasai women discovered that there are traditions that restrict the meals and even the amount of food women eat during pregnancy. Participants in the study indicated that they were not intended to consume “too much” and were advised to avoid foods like meat and beans. According to the findings [26] when such items are eaten, the baby becomes overly huge, causing complications during delivery. Such food restrictions deprive members of a specific cultural group of the essential nutrients included in such ‘forbidden foods’. The findings of the literature study demonstrated that diet choice among pregnant women was subjective to food aversions, economic restrictions, and household food availability [26].

The study’s MUAC measurements of (MUAC < 22 cm) indicates potential malnutrition or nutritional risk at (31.2%). The majority have normal nutritional status (MUAC > 22 cm), accounting for (68.8%). Food insecurity had a negative relationship with the nutritional health of pregnant adolescents, which was similar with the findings of research conducted in Ambo Ethiopia [29] and a global systematic review. Food insecurity, or low intake of nutrients required for optimum nutritional status directly causes undernutrition. The study [30] concluded that hemoglobin levels and anthropometric measurements MUAC determined normal nutritional status and pregnancy outcomes in the majority of pregnant adolescents with nulli and primipara who had large mid-arm circumference and weight gain.

Adolescents having low hemoglobin levels (<11.5 g/dL) were 42, possibly due to iron deficiency or other nutritional concerns, whereas 45 have high hemoglobin levels (>13.5 g/dL). Nutritional and non-nutritional factors can both contribute to anemia. Nutritional anemia is the major cause of anemia worldwide, especially among women of reproductive age and during pregnancy. Thus being comparable to research conducted in Asosa and West Arsi Zones where pregnant women who did not eat fruits were more likely to be anaemic than their counterparts [30].

According to the study (54.9%) of respondents cited distance as a barrier. Antenatal care promotes maternal and child health by providing necessary care, monitoring, interventions during pregnancy, with the objective of maintaining a safe and healthy pregnancy for both the mother and unborn child (54.9%) of respondents cited distance as a barrier [31] According to the findings [32] age, marital status, and the type of employment were all significant predictors of ANC utilization among previously pregnant women. While cohabiting was significantly associated with a higher likelihood of high ANC utilization than single women. Pregnant adolescent of ages (≥15 years) married or cohabitation, and commerce all increased the likelihood of high ANC consumption.

Further, transportation costs, clinical investigation costs, and distance to travelling points may require additional attention, given that approximately (34.7%) of respondents perceived distance to healthcare centers as a barrier to ANC utilization, as well as geographic access to ANC services in Ghana [32]. Moreover, having lack of educational opportunities may contribute to decreased knowledge of ANC and healthcare in general. Additionally, language problems can impede effective communication and comprehension of healthcare information. The survey [33] highlighted perceived variables impacting ANC use, which include poor attitude of health staff, lack of knowledge of ANC services, budgetary restriction and distance to healthcare centers.

The study explores the reasons for not taking the Iron Folic Acid Supplements (IFAS) for 7 days in a week. Among those who took 1 pill, the top reasons mentioned were forgetting 7 respondents, (11.6%), among those who did not receive IFA, ‘no provision’ from the health provider was the most prevalent cause this did concur with the study [34]. Among individuals who received supplements and did not consume them, low palatability, unrecognized need, constipation and forgetfulness were the top causes.

Poor consumption of IFA supplements is a major risk factor for anemia among pregnant women in Sirohi [35]. Though anemia is a complicated and multifaceted condition involving dietary interactions, iron supplementation has the potential to treat a significant amount of anemia in India. As shown in Sirohi, a low intake of IFA supplements as a combination impact of limited inventories and inadequate demand drives anemia among pregnant women. In this study [36] Pregnant women had low adherence to IFAS, and characteristics associated with adherence were knowledge of anemia, knowledge of IFAS, time spent at an ANC visit, number of IFAS pills administered, number of meals, number of children, and distance to the healthcare facility.

### Relationships between Variables

The analysis reviled significant relationship between socio-economic characteristics which include household income, marital status, education, and pregnant adolescent’s nutrition status. Therefore, first hypothesis was rejected, indicating that demographic and socio-economic characteristics having a significant relationship with the nutrition status of pregnant adolescent.

A correlation analysis was conducted to assess the relationship between nutrition knowledge and dietary practices. This suggests that pregnant adolescents with higher nutrition knowledge tend to have more diverse and balanced diets, which is critical for their health and the health of their unborn children. Hence, second hypothesis was rejected, as there was a significant relationship between nutrition knowledge and dietary practices. Enhancing nutrition education for pregnant adolescents could further improve their dietary diversity and overall health.

Furthermore, the analysis revealed a moderate positive correlation between carbohydrate intake and MUAC indicating that higher carbohydrate consumption is associated with better nutrition status. Also, the relationship between vitamin intake and MUAC was strong and statistically significant and therefore, third hypothesis was rejected, as dietary practices and nutrition status had a significant relationship. Improving dietary practices through consumption of a balanced variety of foods could enhance the nutrition status of pregnant adolescents.

Pregnant adolescents who had higher levels of nutrition knowledge were more likely to regularly consume IFAS, which is critical for preventing anemia and ensuring proper fetal development. The analysis also showed that adolescents who understood the benefits of IFAS were more compliant with their intake, compared to those who lacked knowledge therefore fourth hypothesis was rejected, confirming that there was significant relationship between the consumption of IFAS and nutrition knowledge. This calls for more targeted education programs to enhance IFAS compliance and, subsequently, improve maternal health outcomes.

## Conclusions

The study found that majority of pregnant have dropped out of school owing to pregnancy. This study found that bulk of the pregnant adolescent division is in middle adolescence and comes from relatively poor homes that rely mostly on livestock farming, selling, and purchasing food. Interventions that boost and diversify food crop production in these dry areas are extremely necessary to make food more accessible.

Pregnant adolescents in this study have harmful dietary patterns such as meal skipping, lack of diversification and low nutrient intake. In the study adolescent pregnancy were found to have both undernutrition and micronutrient deficiencies. This necessitates interventions that address the special nutritional demands of pregnant adolescent at this key developmental stage, promoting healthy growth while reducing the risk of health problems.

The nutrition status of pregnant adolescent is low as stated by MUAC and relatively narrow range of haemoglobin readings, with most falling within the lower normal range. Thus low nutritional status being harmful to their health. The nutrition status was impacted by the meals consumed and amount of kilocalories ingested. The nutritional status of pregnant adolescents in this community is low. This is due to drought, which affects livestock. Additionally, existence of animals rarely convert to money to buy food. Low nutrition status was linked to insufficient dietary consumption. Low diet eating patterns were caused by a lack of education and nutrition understanding, combined with a lack of sufficient means to purchase food.

This study concludes that lack of ANC visits was linked to low nutrition education and thus lack of IFAS consumption. On the other hand, linked to an increased risk of unfavorable outcomes such as premature lab our, low birth weight, and perinatal death. Thus the need for nutrition education.

## Abbreviations

ANC: Antenatal Clinic
ASAL: Arid and Semi-Arid Land
Hb: Hemoglobin
MUAC: Mid-upper Arm Circumference
RDA: Ready Dietary Allowance

## Acknowledgements

The authors are grateful to Almighty God, Kenyatta University, Food nutrition and dietetics department for their professional direction, counsel, and support throughout my studies, County Government of Kajiado, through the County Directors of Health and Education, for enabling me to collect data, research assistants, and community health promoters and to my family members for their efforts, encouragement, and moral support.

## Data Availability Statement

The data supporting the outcome of the research work has been reported in this manuscript.

## Author Contributions

Dr. Joseph Kobia: Supervision, Writing- review & editing

Dr. Dorcus Mbithe David-Kigaru Writing- review & editing

